# DRIVE-KG: Enhancing variant-phenotype association discovery in understudied complex diseases using heterogeneous knowledge graphs

**DOI:** 10.1101/2025.08.19.25333942

**Authors:** Ananya Rajagopalan, Tram Anh Nguyen, Lindsay A. Guare, Andre Luis Garao Rico, Rasika Venkatesh, Lannawill Caruth, Regeneron Genetics Center, Penn Medicine BioBank, Anurag Verma, Marylyn D. Ritchie, Molly A. Hall, Joseph D. Romano, Shefali Setia-Verma

## Abstract

Multi-omics data are instrumental in obtaining a comprehensive picture of complex biological systems. This is particularly useful for women’s health conditions, such as endometriosis which has been historically understudied despite having a high prevalence (around 10% of women of reproductive age). Subsequently, endometriosis has limited genetic characterization: current genome-wide association studies explain only 11% of its 47% total estimated heritability. Graph representations provide an intuitive and meaningful way to relate concepts across diverse data sources and address fundamental sparsity and dimensionality challenges with multi-omics data analysis. Here we present DRIVE-KG (Disease Risk Inference and Variant Exploration-Knowledge Graph), which uses a heterogeneous graph representation to integrate biological data from multi-omics datasets: dbSNP, NCBI Human Gene, Omics Pred, GTEx, and Open Targets. We drew directly from the knowledge captured in these data, using nodes to represent genes, single nucleotide polymorphisms, proteins, and phenotypes, and edges to represent relationships between these concepts. We trained two models using DRIVE-KG: a link prediction model to suggest associations between SNPs and two pilot phenotypes (endometriosis and obesity), and a graph convolutional network (GCN) to classify patient-level endometriosis status. We conducted the patient-level classification using data from 1,441 Penn Medicine BioBank participants with gold standard chart-reviewed endometriosis status. The link prediction model uncovered 66 high-confidence (score ≥ 0.95) previously unreported SNP-endometriosis associations. Many of these variants were linked to obesity/body mass index traits (24.2%), lipid metabolism (6%), and depressive disorders (4.5%), showing agreement with emerging hypotheses about endometriosis etiology. In contrast, 11% of the 149 high confidence, candidate SNP-obesity associations (score ≥ 0.9888) were in LD with known obesity associations. The GCN to classify patient endometriosis status had an AUPRC of 0.738 compared to 0.679 for a genetic risk score. Despite this moderate improvement, we found that the GCN learned meaningful stratification of underlying adenomyosis signal and severe grades of endometriosis. We have demonstrated that heterogeneous integration of multi-omics data is valuable for diverse downstream tasks—including discovery and clinical prediction—particularly for understudied diseases where traditional genomic approaches are insufficient.

## 1. Introduction

Multi-omics data integration involves combining diverse biological data types—including genomic, proteomic, epigenomic, and metabolomic data—to provide comprehensive insights into the mechanisms of diseases.^1^ This approach is particularly useful for uncovering complex molecular interactions and biological pathways that remain hidden when analyzing individual data types in isolation, and leads to a more complete understanding of disease etiology and progression.^2^ Despite this potential, significant computational challenges exist with multi-omics data integration. The high-dimensional, heterogeneous, and sparse nature of multi-omics data creates substantial barriers in developing analysis methods that are both biologically interpretable and computationally efficient.^3^ These integration challenges highlight the critical need for innovative analytical frameworks that can effectively capture and represent the complex relationships inherent in multi-omics data.

Knowledge graphs have emerged as a promising solution to address these multi-omics integration challenges. By representing biological entities as nodes and the known relationships that exist between them as edges, knowledge graphs provide a framework that can capture the complex, interconnected nature of biological systems across data modalities.^3^ Graph-based approaches are particularly well-suited for multi-omics data because they can naturally accommodate heterogeneous data types while preserving their semantic relationships and biological context. Given large-scale data of biological knowledge from different sources, graph representations allow for a natural, semantically meaningful way to encode the relationships within and between data. For multi-omics integration, a heterogeneous knowledge graph representation offers particular advantages.^4^ Unlike homogeneous graphs where all nodes and edges represent the same type of entity or relationship, heterogeneous knowledge graphs can simultaneously represent multiple entity types (e.g., genes, proteins, metabolites) and diverse relationship types (e.g., regulatory interactions, protein-protein interactions, metabolic pathways) within a single unified structure.^3^ Another major advantage of knowledge graph-based approaches for multi-omics integration is that they do not require data from the same individuals across different -omics layers. Instead, these methods can leverage population-level patterns and biological knowledge to create comprehensive representations that integrate information from disparate studies and cohorts. Existing biological knowledge encoded in the graph structure can then be leveraged via network analyses for a variety of purposes: making use of existing relationships in the graph to predict new connections, or for downstream tasks such as patient-level classification.

For many well-characterized complex diseases like obesity, extensive genetic studies have successfully explained a substantial proportion of disease heritability. Furthermore, polygenic risk scores (PRS) demonstrate strong predictive performance; for obesity, typical AUC values for PRS are around 0.8.^5^ These genetic studies are enabled by large-scale biobanks (e.g., UK Biobank,^6^ Penn Medicine BioBank,^7^ and All of Us^8^) which are collections of de-identified genetic, lifestyle, clinical and biological data. In contrast, endometriosis—one of many complex gynecological conditions—affects approximately 10% of reproductive age women yet remains severely understudied and underfunded.^9^ Due to its consequent underdiagnosis, traditional genomic approaches fall short as there is a much lower prevalence observed in biobanks. Despite a heritability estimate of 47%,^10^ variants identified through GWAS studies of endometriosis to-date have only been able to explain roughly 11%^11^ of this heritability, and the predictive accuracy of genetic risk scores have been inconsistent as a result.^12^ The ability of knowledge graph methods to integrate information across disparate studies and populations, without requiring matched individual-level data, makes them especially suited to address these gaps. While previous studies have applied graph representations to integrate multi-omics for disease discovery^13^ and prediction,^14^ we are not aware of effective applications of these methods to complex and understudied conditions such as those in women’s health. By leveraging existing multi-omics data from independent studies and incorporating established biological relationships contributing to disease, these approaches can potentially uncover disease mechanisms and improve predictive models for conditions where traditional large-scale genomic studies have been insufficient.

This study presents an innovative framework for integrating multi-omics data about disease associations through a heterogeneous knowledge graph representation. We hypothesize that by systematically incorporating knowledge from diverse -omics data sources within a unified graph structure, we can identify previously hidden candidate relationships that are both biologically meaningful and mechanistically relevant to disease pathogenesis. Combining large-scale biological data in this way will improve our ability to discover meaningful and interpretable insights about disease pathogenesis. This is particularly impactful for historically understudied women’s health conditions where traditional genomic methods have not fully captured the complexity of disease etiology.

## 2. Methods

### 2.1. Knowledge Graph Data Sources

To construct a heterogeneous knowledge graph (KG), we integrated multiple biological data sources (node and edge types summarized in Table 1). The graph is comprised of four primary node types: single nucleotide polymorphisms (SNPs), genes, proteins, and phenotypes.

**Table 1.** Knowledge Graph Data Sources.

| Graph Concept | Data Source | Features |
| --- | --- | --- |
| Node: SNP | dbSNP<sup>15</sup> | Bulk RefSNP JSON files |
| Node: Phenotype | Human Phenotype Ontology<sup>16</sup> | HP Ontology |
| Node: Gene | NCBI Gene<sup>17</sup> | Homo Sapiens, filtered to 9606 Taxa ID |
| Node: Protein | UniProtKB<sup>18</sup> | Swiss-Prot Entries (XML) |
| Edge: SNP to Gene | GTEx<sup>19</sup> | version 8, eQTL |
| Edge: SNP to Phenotype | Open Targets<sup>20</sup> | GWAS Summary Statistics |
| Edge: Gene to Phenotype | OmicsPred<sup>21</sup> | PheWAS Summary Statistics |
| Edge: SNP to Protein | OmicsPred<sup>21</sup> | Olink, Somalogic (Proteomics) |
| Edge: Gene to Protein | UniProtKB<sup>18</sup> | Swiss-Prot Entries (XML) |

#### 2.1.1. Node Data Sources

We derived SNP nodes from the Single Nucleotide Polymorphism Database (dbSNP).^15^ To avoid node type imbalance caused by including tens of millions of variants, we included only SNPs with minor allele frequency (MAF) *≥* 0.01 in the Penn Medicine BioBank (PMBB). We obtained phenotype nodes from the Human Phenotype Ontology (HPO),^16^ providing standardized disease and trait terminology. We sourced gene information from the National Center for Biotechnology Information (NCBI) gene database,^17^ and derived protein nodes from the Universal Protein Resource (UniProt).^18^

#### 2.1.2. Edge Data Sources

We incorporated five distinct edge types connecting the four biological entities (SNP, gene, phenotype, and protein) into the KG. We established SNP-to-gene associations using expression quantitative trait loci (eQTL) data from version 8 of the Genotype-Tissue Expression (GTEx) project.^19^ We derived SNP-to-phenotype relationships from Open Targets,^20^ which aggregates evidence from genome-wide association studies (GWAS) and other sources. We obtained gene-to-phenotype associations from PheWAS (phenome-wide association study) summary statistics, published from OmicsPred.^21^ We established SNP-to-protein connections through OmicsPred, and sourced gene-to-protein relationships from UniProt annotations.^18^

### 2.2. Disease Risk Inference and Variant Exploration-Knowledge Graph (DRIVE-KG) Construction

When designing the DRIVE-KG schema (Figure 1), we prioritized having dense node properties and fewer edge types to avoid sparsity issues when performing machine learning analyses on the graph. We maintained cross references of node type identifiers across databases (‘dbXrefs’) as properties of the gene and phenotype node for possible future querying by other researchers. The ‘tissue expression’ property of the ‘hasEqtl’ edge is an array of length 53 (number of tissues) that contains all tissue-specific beta values for eGenes in this dataset. Since eGenes (a gene whose expression is influenced by eQTLs) are tissue-specific in the Omics Pred dataset, we aggregated all tissue-specific effects for a particular SNP–Gene pair into this list. If a particular SNP–Gene pair did not have a tissue-specific association, the beta value for that entry was set to 0.

**Figure 1.**
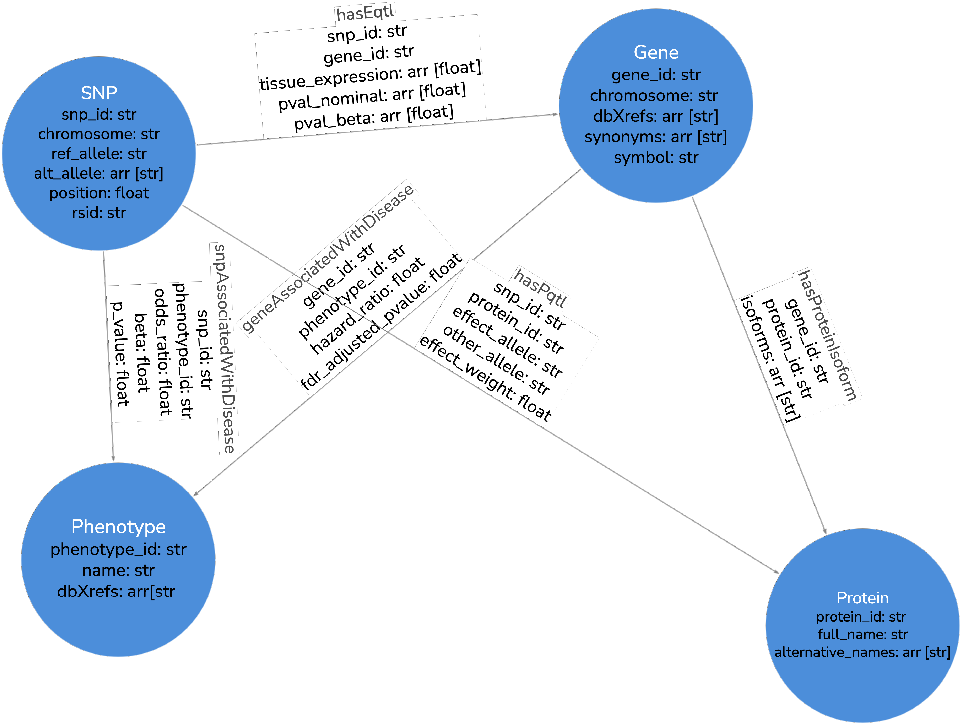
Knowledge Graph Schema.

Omics Pred pQTl data pertain to the same 53 tissue types as published by the GTEx consortium, with the exception of Bladder, Fallopian tube, and Cervix (Endocervix & Ectocervix), resulting in a total of 49 tissue types. The tissue expression array was mapped in the same alphabetical order as used by the GTEx consortium,^19^ so the index of a given beta value in this array can be used to infer from which tissue it came from.

The DRIVE-KG construction process involved several key steps: first, we standardized identifiers across all data sources (e.g., harmonized to the same Entrez identifier for all gene-related data and ensured that hg38 was used to represent all SNP-related information). Finally, we ingested these files into Memgraph^22^ for subsequent visualization and analysis.

For the purposes of model training, we removed all unconnected nodes (node with degree = 0) in the graph (removing 94.2% of the original 13,806,079 nodes), yielding a total of 801,112 nodes and 1,390,440 edges in the graph. The removed nodes consisted mainly of highly specific genetic variants and phenotypes without documented associations in our data sources. This pre-processing step preserved the core connectivity structure necessary for effective graph-based learning and link prediction analyses.

### 2.3. Link Prediction

To identify previously unreported, biologically relevant relationships within DRIVE-KG, we employed link prediction methods that leverage our multi-modal data structure. The core hypothesis underlying our approach is that missing connections (edges) between biological entities can be predicted by exploiting the rich connectivity patterns and semantic relationships encoded in the existing graph structure. Our link prediction framework operates on the principle that biologically meaningful associations often exhibit characteristic graph topology patterns. For instance, if a SNP is associated with multiple genes that are collectively linked to a specific phenotype, this suggests a potential direct SNP-phenotype relationship that may not be explicitly captured in current databases. Similarly, proteins sharing common genetic variants and phenotypic associations may indicate previously uncharacterized protein-protein interactions or shared functional pathways.

In our case, we were interested in predicting whether there should be a link between a given SNP node and two pilot phenotypes: endometriosis and obesity, both complex traits with contrasting levels of previous study and heritability. We chose obesity as a comparison phenotype given its extensive genetic characterization compared to endometriosis. Consequently, there are a large number of existing genome-wide significant variants in association with obesity for the link prediction model to train on (112 compared to 67 for endometriosis).

We obtained 64-dimension node embeddings using Memgraph’s node2vec^23^ module, and then used these embeddings as input for a pairwise multilayer perceptron (MLP) model developed using the PyTorch Lightning^24^ module. The MLP architecture is a three-layer feed-forward network, with rectified linear unit (ReLU) activation functions and dropout after the first two linear layers. The MLP input of 64-dimension embeddings are projected using the first linear layer to 128-dimensions. These embeddings are sequentially reduced with each of the subsequent two linear layers (from 128-dimension to 64-dimension, then from 64-dimension to 1-dimension). The final linear layer of the model produces a scalar score, indicating the ‘probability’ that a particular SNP-phenotype edge should exist.

We conducted link prediction of snpAssociatedWithDisease edges with Memgraph’s native link prediction module with a training-validation split ratio of 0.7. We queried the link prediction model for associations with this phenotype using their HPO IDs (HP 0030127 for endometriosis and HP 0001513 for obesity). We explored the model’s output values for each phenotype (transformed to 0–1 by the final sigmoid activation function), indicating the model’s confidence in a particular edge. In order to assess the sensibility of the model’s predictions, we cross-referenced predicted associations against existing GWAS results and explored the neighborhood of high-confidence candidate associations in DRIVE-KG.

### 2.4. Endometriosis Patient Classification

In order to assess the utility of DRIVE-KG for disease risk stratification, we trained a graph convolutional network. In particular, we explored the potential of this graph to accurately stratify disease risk at the patient level when applied to endometriosis. We obtained chart reviews for a cohort of 1,441 patients from the PMBB^7^ with their status of endometriosis or adenomyosis (case or control) labeled, as these conditions are often grouped together during phenotyping despite likely differences in their underlying pathogenesis. For endometriosis cases, the chart review also documented surgical disease stage according to established clinical classification systems, enabling both binary case-control analyses and stage-specific investigations. Additional demographics for this cohort are in Table 2.

**Table 2.** Demographics of PMBB Chart-Reviewed Cohort.

| Characteristic | Category | Overall (n=1441) | Control (n=732) | Case (n=709) |
| --- | --- | --- | --- | --- |
| **Sequenced gender** | Female | 1441 (100.0%) | 732 (100.0%) | 709 (100.0%) |
|  | Male | 0 (0.0%) | 0 (0.0%) | 0 (0.0%) |
| **Ancestry group** | AFR | 613 (42.5%) | 256 (35.0%) | 357 (50.4%) |
|  | AMR | 30 (2.1%) | 20 (2.7%) | 10 (1.4%) |
|  | EAS | 20 (1.4%) | 13 (1.8%) | 7 (1.0%) |
|  | EUR | 746 (51.8%) | 422 (57.7%) | 324 (45.7%) |
|  | SAS | 30 (2.1%) | 19 (2.6%) | 11 (1.6%) |
|  | UNKNOWN | 2 (0.1%) | 2 (0.3%) | — |

To curate the graph classification dataset, we generated per-patient copies of DRIVE-KG from the chart-reviewed cohort. We did this by first exporting the base KG and graph connectivity information from Memgraph into Python (PyTorch Geometric library^25^). Then, we assigned the value of each SNP node, for each patient’s graph, as the imputed genotype dosage (where the value indicates the dosage of the alternate allele at a given locus) from the imputed genotyping data in the 2024 PMBB data release 3.0, which was processed using PLINK2.0.^26^ Here, the prediction task is whether the entire graph represents endometriosis/adenomyosis (case) or not (control). For all non-SNP node types, we assigned 8-dimension embeddings learned using the node2vec module. We also added per-patient covariates (age and the first 5 of overall ancestry principal components (PCs)) to the classifier.

We leveraged PyTorch Lightning to load the patient graph objects and train the graph classifier. The graph classifier architecture contains two convolutional layers for all edge types, with SAGEConv layers wrapping each of the edges (we chose SAGEConv due to its inductive capabilities, and ability to efficiently scale to large graphs). We used 64-dimension projections of the 8-dimension embeddings learned on all nodes as model input. We used ReLU activation functions for the two convolutional layers, and the ‘sum’ aggregation function (which generates 64-dimension tensors for each node type). We then generated pooled GNN embeddings over the SNP node type, which creates 64-dimension tensors for the entire graph. We pooled over the SNP node type alone since only the SNP node values differ across patient graphs; therefore, it is likely to contain biological signal that can be used to distinguish patients. We concatenated the pooled, 64-dimension tensors with the per-patient covariates data (6-dimension for age and 5 PCs) to get 70-dimension tensors. We fed these tensors into the first linear layer, which transformed them into 64-dimension tensors with a ReLU activation function. Finally, the second linear layer contained a sigmoid activation function that generated a binary classification value (single probability per graph/patient) from the 64-dimension tensors. For each epoch, we specified a batch size of 16 and a learning rate of 0.001 for the optimization function.

We compared the performance of disease risk prediction using DRIVE-KG to a genetic risk score (GRS)^11^ developed and evaluated on the same patient cohort (with the same training-validation split of patients between DRIVE-KG and the GRS). We calculate the GRS by linearly combining the patient’s genetic information alone (no additional data).

## 3. Results

### 3.1. Link Prediction

We investigated link predictions for two phenotypes of interest: endometriosis (HP 0030127) and obesity (HP 0001513). For each phenotype, we obtained recommendations from the model of SNP-phenotype links, ranked by their binary classification score and sorted by whether they are existing or candidate associations.

#### 3.1.1. Endometriosis

Our link prediction analysis identified 66 high confidence, previously unreported SNP-endometriosis associations (model score *≥* 0.95). Notably, many of these predicted associations involved SNPs with well-established links to other complex traits and conditions, yet had not been previously associated with endometriosis in the literature (except for rs10828249 located on *MLLT10* ^27^). Specifically, 24.2% of the top-ranked predicted SNP-endometriosis links involved variants previously associated with body mass index (BMI)/obesity-related traits, 6% with lipid metabolism (triglycerides and HDL), and 4.5% with depressive disorders. Figure 2 compares all existing vs. candidate SNP-endometriosis associations in DRIVE-KG with respect to model confidence. We observed a higher confidence for new, candidate associations compared to existing association scores. This could be because the model is undertrained on endometriosis associations compared to more well-studied diseases (i.e., those with a greater number of genome-wide significant associations) and biased towards positive predictions as a result. There were 67 existing SNP-endometriosis associations for the link prediction model to train on, which had a mean confidence of 0.635.

**Figure 2.**
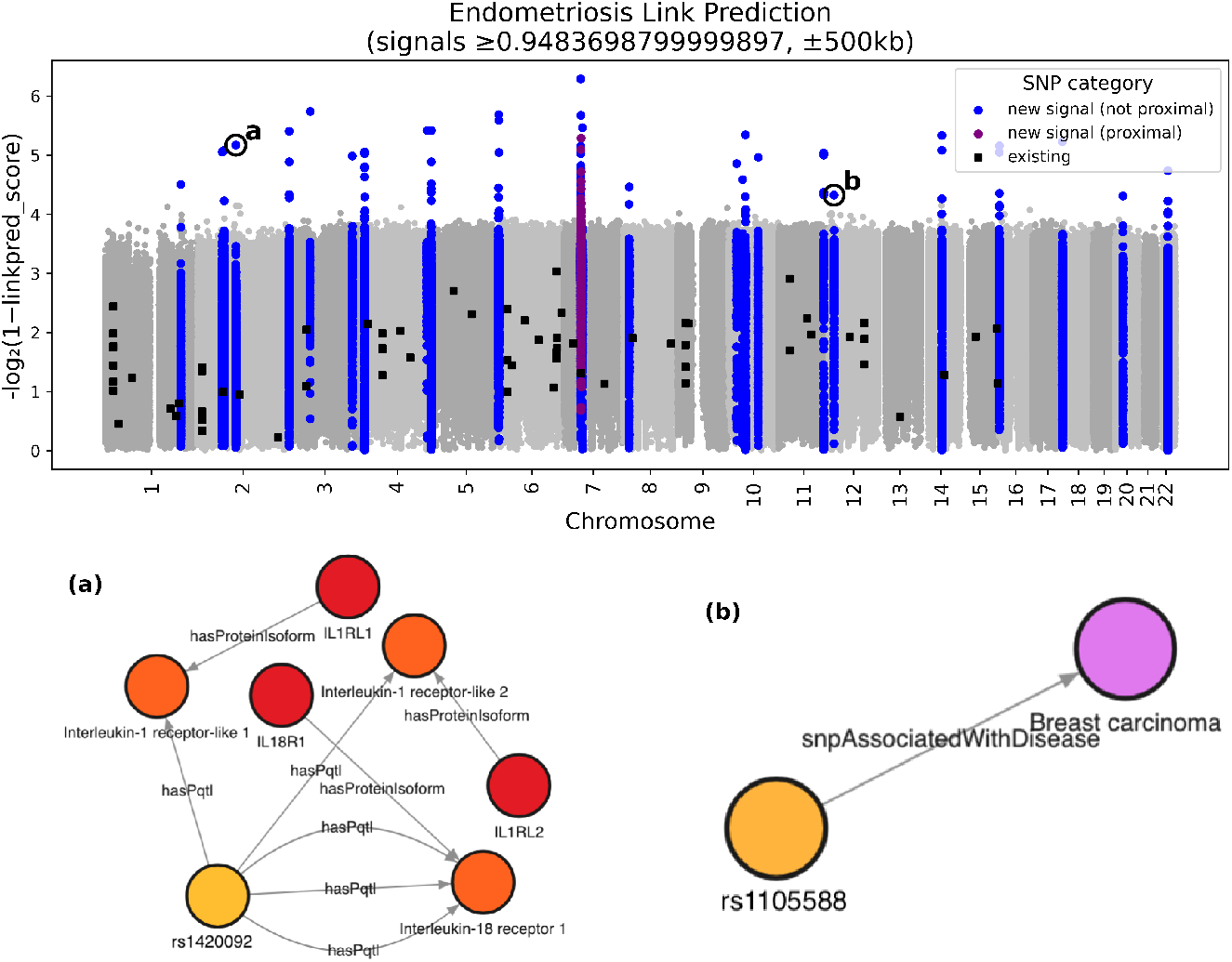
Endometriosis Link Prediction: comparison of new and existing signal with respect to model confidence.

#### 3.1.2. Obesity

In contrast to the endometriosis results, link prediction for obesity yielded markedly different patterns, reflecting the extensive existing knowledge for these well-characterized phenotypes. There were 112 existing SNP-obesity associations for the link prediction model to train on, which were ranked with a mean confidence of 0.926 (much higher than the corresponding mean confidence of 0.635 for endometriosis). Moreover, the confidence scores for predicted candidate obesity associations were substantially lower (mean score = 0.276) compared to candidate endometriosis predictions (mean score = 0.758). Notably, 11% of the top-ranked candidate SNP-obesity associations (n = 149, score *≥* 0.9888) were predominantly variants in linkage disequilibrium (LD) with previously established obesity-associated loci (e.g., genes *NEGR1* ^28^ and *DNAJC27-AS1* ^29^) rather than unreported associations. This suggests that the predicted associations largely recapitulated existing knowledge of metabolic pathways and obesity-related biological mechanisms already well-documented in the literature. The network visualizations for the high-confidence, candidate SNP-obesity associations in Figure 3 (rs74432706 and rs329642) demonstrate that these predictions are primarily driven by neighboring connections to obesity phenotypes, as opposed to distinct biological signals.

**Figure 3.**
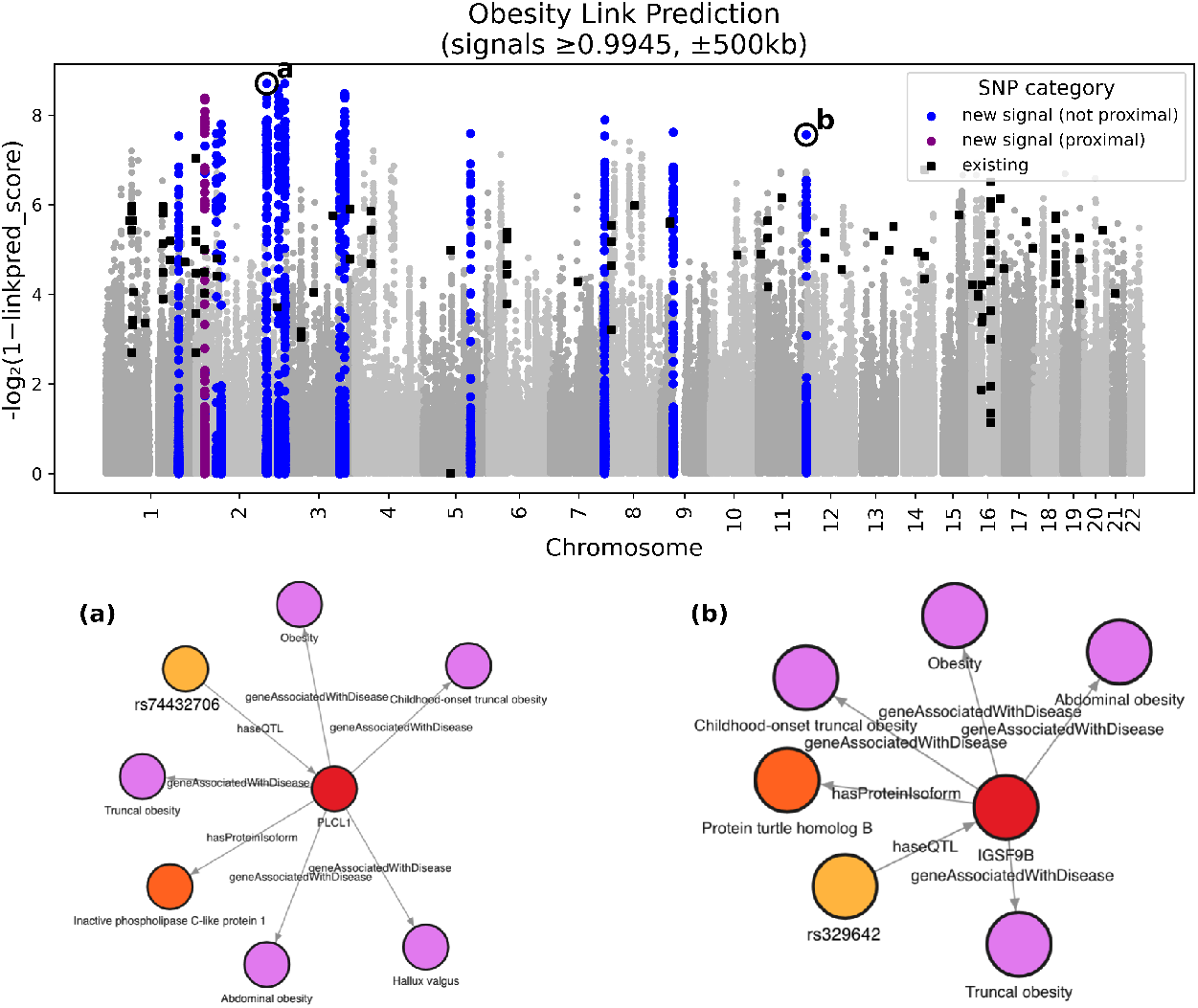
Obesity Link Prediction: comparison of new and existing signal with respect to model confidence.

### 3.2. Endometriosis Patient Classification

Our cohort consisted of 1,411 PMBB participants assigned female at birth with imputed genotyping data and a chart reviewed label (0 or 1) for having either endometriosis/adenomyosis or not. With a 70-30 training-validation split, the graph classifier had an AUPRC of 0.738 and a loss of 0.75. We compared this to a GRS developed and evaluated on the same patient cohort, which had an AUPRC of 0.679 (additional metrics displayed in Table 3). Since this is a binary classification task, the model provides a score (value between 0 and 1, inclusive) for its predicted likelihood of a given patient having either an endometriosis or adenomyosis diagnosis. Figure 4 provides model score distributions stratified both by disease subtypes from chart reviews (Figure 4a) and by surgically confirmed endometriosis stages (1-4) (Figure 4b).

**Table 3.** Patient Classification Evaluation (Mean *±* Standard Deviation)

| Model Type | AUC-ROC | AUPRC | F<sub>1</sub> | Accuracy |
| --- | --- | --- | --- | --- |
| DRIVE-KG | 0.569 ± 0.032 | 0.738 ± 0.033 | 0.415 ± 0.028 | 0.420 ± 0.024 |
| GRS | 0.476 ± 0.033 | 0.679 ± 0.031 | 0.007 ± 0.006 | 0.296 ± 0.022 |

**Figure 4.**
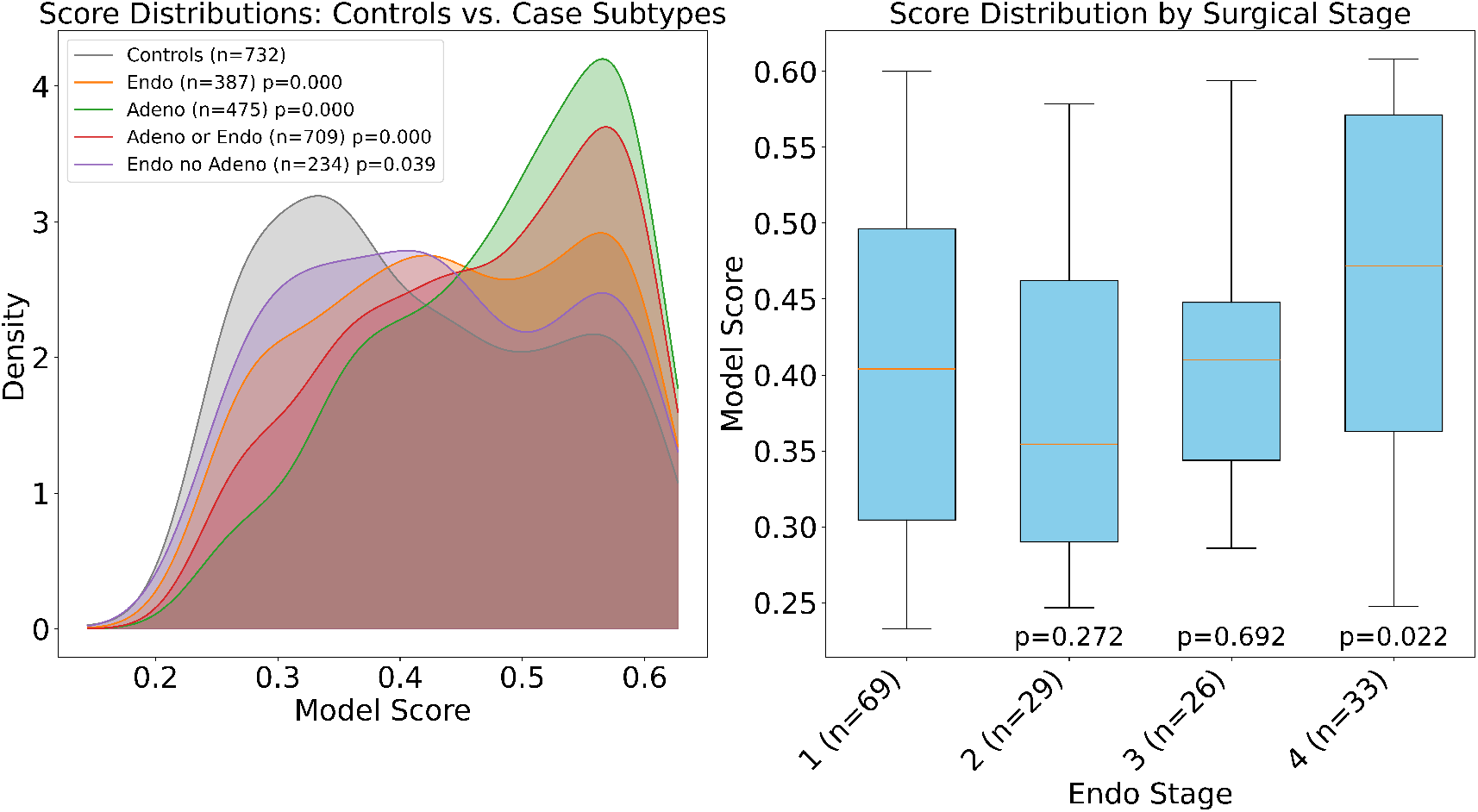
(a) Model score distribution for chart-reviewed disease subtypes; (b) Model score distribution by chart-reviewed, surgically confirmed endometriosis stages.

## 4. Discussion

This study demonstrates the potential of heterogeneous KG approaches to advance our understanding of understudied diseases where traditional genomic methods have reached their current limits. Our findings provide several key insights into the utility of integrating multiomics data through graph-based representations for both discovery and clinical prediction applications. The link prediction results reveal a striking contrast between well-studied and understudied phenotypes, validating our central hypothesis. For endometriosis, our approach successfully identified high-confidence candidate SNP associations involving variants previously linked to BMI, lipid metabolism, and depressive disorders—yet predominantly not previously associated with endometriosis itself. Additionally, visualizing the network of high-confidence SNP-endometriosis associations revealed connections to underlying inflammatory and cell proliferation pathways, providing additional insight into shared biological mechanisms. This pattern suggests shared biological pathways between endometriosis and these comorbid conditions, which aligns with growing clinical evidence of their frequent co-occurrence.^30–32^ The identification of metabolic and behavioral trait variants as endometriosis candidates highlights emerging mechanistic hypotheses^11,27,33^ about disease etiology, potentially explaining some of the substantial “missing heritability” gap where traditional polygenic risk scores capture only 11% of the estimated 47% heritability.^11^ Conversely, for obesity—a trait with extensive genomic characterization—our link prediction yielded lower confidence scores and predominantly identified variants in linkage disequilibrium with known associations. This outcome validates our approach: DRIVE-KG appropriately finds fewer novel discoveries for well-mapped biological landscapes while successfully uncovering genuine candidates for understudied conditions.

Our patient classification for endometriosis results demonstrates modest but meaningful improvements over traditional genetic approaches. The DRIVE-KG-based model achieved an AUPRC of 0.738 compared to 0.679 for the GRS alone. These moderate effect sizes show potential clinical relevance for a condition where existing genomic tools perform poorly: the model showed enhanced prediction for adenomyosis and demonstrated particular strength in identifying patients with severe stages of endometriosis (stage 4). The improved performance likely stems from DRIVE-KG’s ability to capture complex multi-omics relationships that single-modality approaches miss, even when the only patient-specific input is genetic data. Rather than relying solely on individual genetic variants, our framework leverages the broader biological knowledge encoded in protein interactions, gene expression patterns, and phenotypic associations. This integrated approach may be particularly valuable for conditions like endometriosis where the genetic architecture is complex and poorly understood.

These findings have broader implications for addressing healthcare disparities in women’s health research. Endometriosis exemplifies the challenges facing many gynecological conditions: high prevalence (affecting around 10% of women of reproductive-age^9^), significant clinical impact, yet persistent research neglect leading to limited therapeutic options. Our results suggest that graph-based approaches can help bridge these knowledge gaps by extracting additional insights from existing multi-omics data without requiring massive new cohort studies that may be infeasible for understudied populations. The identification of shared pathways with well-studied traits (BMI, depression) also suggests opportunities for re-purposing existing therapeutic targets and biomarkers. If endometriosis shares mechanistic pathways with metabolic or psychiatric conditions, this could accelerate drug development and improve patient care through better understanding of comorbidity patterns.

Our study has several limitations. First, the construction of DRIVE-KG relied on existing databases that may contain biases toward well-studied biological pathways, potentially limiting discovery of entirely novel mechanisms. Second, we account for harmonizing variants across major and minor allele when creating DRIVE-KG by checking for SNP identifier matches across both directions of alleles; however, we do not account for differences in the effect allele when processing the PMBB genotype data to make patient-specific graphs, and simply drop variants that are in PMBB data but not in DRIVE-KG. Changing the SNP node representation in the future to be per alternate allele would enable us to more completely utilize the genotype data when using the DRIVE-KG representation for patient classification. Third, we filtered the SNP nodes at a MAF *≥* 0.01 during training. This was to reduce memory constraints from Memgraph, which relies on an in-memory database and storage, but this restricted our analysis to common variants. In the future, we aim to incorporate rare variants as well (not filtered at MAF 0.01) for exploring associations with higher effect sizes. For patient classification, our modest AUC improvements—while significant—will require further validation in independent cohorts. Furthermore, since we performed patient classification on a single-institution cohort, generalizability across diverse populations remains to be established. Lastly, we emphasize that the development of DRIVE-KG is ongoing, and this paper is meant to highlight the first iteration of features that can support meaningful biomedical discovery. It is critical for our KG creation to be structured in a way that supports continual incorporation and refreshing of data from its source databases. While this has not been achieved in the current iteration of DRIVE-KG, we plan to address this in subsequent development.

Our future work will focus on incorporating additional data modalities (e.g., transcriptomics and metabolomics) to further enhance the comprehensiveness of DRIVE-KG. Additionally, experimental validation of our predicted SNP-endometriosis associations will be crucial for translating these computational findings into biological insights. Finally, extending this framework to other understudied conditions and long-term outcomes could help address broader research inequities.

In conclusion, our study demonstrates that DRIVE-KG offers a promising approach to advance precision medicine in understudied diseases. By integrating multi-omics data in a unified framework, we identified previously unreported biological candidates for endometriosis (that underscore emerging hypotheses about disease mechanisms) and achieved improved patient classification compared to traditional genomic approaches. These findings highlight the particular value of graph-based methods for conditions where conventional genomic tools have reached their current limits, offering a path forward to address knowledge gaps in understudied populations.

## Data Availability

All data produced in the present study are available upon reasonable request to the authors. All code used in this work is available at https://github.com/Setia-Verma-Lab/multi_omics_kg.

## 5. Acknowledgments

We gratefully acknowledge the PMBB for providing data and thank the patient-participants of Penn Medicine who consented to participate in this research program. The PMBB is approved under IRB protocol #813913 and #852155 and supported by Perelman School of Medicine at University of Pennsylvania, a gift from the Smilow family, and the National Center for Advancing Translational Sciences of the National Institutes of Health under CTSA award number UL1TR001878. The research reported in this publication was supported by the Eunice Kennedy Shriver National Institute of Child Health and Human Development of the National Institutes of Health under award number R01HD110567-01A0, and by the US National Library of Medicine under award number R00LM013646.

## Code Availability

All code used in this work is available at https://github.com/Setia-Verma-Lab/multi_omics_kg

